# Efficacy of smoking cessation in spirometry results of COPD smokers: a randomized controlled clinical trial

**DOI:** 10.1101/2020.04.29.20085894

**Authors:** abbas alipour, mehran zarghami, ali sharifpour, fatemeh taghizadeh

## Abstract

**Background:** Nicotine replacement therapy (NRT) may be more effective if it is combined with short cognitive-behavioral interventions for smoking cessation in chronic obstructive pulmonary diseasesmokers.

**Material and methods:** To examine the effectiveness of guided self-change (GSC), in a randomized controlled clinical trial, 57 men ranging from 45 to 77 years old were randomly assigned to three 19-member groups (GSC, NRT, and combined GSC-NRT).

The primary data on smoking cessation and pulmonary functions were examined during 29 weeks using General Linear (GEE) Model status, intention-to-treat analysis, and repeated measures ANOVA test.

**Results:** A total of 9 (47.4%) of the participants in the GSC and combined groups and 4 (21.1%) participants in the NRT group reported *total abstinence rate* from smoking by the end of 29 weeks. *Daily cigarette* number was changed from 24 to 4 in GSC group, 26 to 11 in NRT group, and 20 to 6 in combined group. The GEE model revealed that this variable decreased in GSC group more than two other groups significantly (P=0.003). Moreover, the FVC level of the NRT group was lower than the GSC group (P=0.04), and the FEV in the NRT group was lower than GSC group (P=0.02). Furthermore, the level of FEV1/FVC act/pred in the NRT group was lower than GSC group (−6, 95% CI: -10.4-(−1.5), P=0.008) and it was also lower in the combined group than the GSC group (−6, 95% CI: -11.3-(−0.5), PV=0.03).

**Conclusion:** GSC and combined GSC-NRT treatments were equally effective in abstinence rate. Moreover *Daily cigarette* and the FEV1/FVC act/pred in GSC group was more than two other groups, indicating the health professionals can apply GSC alone in smoking cessation and improve lung function of COPD smokers.

## Introduction

Almost one-third of the adult people in industrial countries and seventy percent in countries of Asian are smokers, and two-thirds of lifelong smokers eventually die as a result of smoking(1). Smoking is the main reason of chronic obstructive pulmonary disease (COPD) (2, 3). COPD affects approximately 10% of the global population and is growing in prevalence annually (4). It affects almost 400 million people (5) and is the third leading cause of death worldwide (6). COPD is characterized by persistent respiratory symptoms and chronic airflow limitation and is associated with comorbidities (7) and progressive non-reversible narrowing of airways, and is mainly caused by cigarette smoking (4).The natural history of COPD demonstrates that FEV1 rapidly declines in smokers who are susceptible to cigarette smoke(8). The susceptible smokers who quit smoking not only recover a little but also the rate of the FEV1 drop is no longer steep (8, 9). Pulmonary function tests are recommended to measure FEV1 in all smokers (9). The use of FEV1/FVC is a traditional amount of obstruction in airways to detect airways obstruction during spirometry testing (10). Smoking quitting is the most effective idea for reducing the progress of the COPD and decreasing mortality in nearly fifty percent of smokers with COPD (2, 11–13). Use of nicotine replacement therapy (NRT) in combination with counseling have proven to help these patients to quit (4, 11) and have efficacy in smoking quitting rates improvement. When combined with a brief counseling, NRT has been effective in increasing smoking cessation and persistent abstinence in COPD smokers (12). One approach in this regard is to combine guided self-change with NRT (12).

**Guided self-change (GSC)** is a brief cognitive-behavioral intervention that was first designed to help alcoholics to identify and develop their abilities to solve their dependency problems (15). This culture-sensitive treatment approach has been applied successfully to a variety of individuals, couples, or groups with problematic alcohol/cigarette/substance abuse (14–20). Since GSC has not been compared with NRT in smoking cessation yet, the investigators considered a randomized, controlled clinical trial to compare the efficiency of them for cigarette smoking cessation in COPD patients in Iranian culture.

## Materials and Methods

This trial was done in the Psychiatry and Behavioral Sciences Research Center in Addiction Institute and Lung Research Center of Mazandaran University of Medical Sciences in Sari, Iran. The trial protocol was approved by Ethics committee of Mazandaran University of medical sciences (IR.MAZUMS.REC.95.2137), registered in the Iranian Registry of Clinical Trials (IRCT201609271457N11; www.irct.ir), and executed in accordance with the Declaration of Helsinki and its subsequent revisions. Subjects were informed of the research protocol and their right to withdraw from the trial at any time and then they provided a written informed consent. The study was performed from December 2016 to November 2017. The statistical population included all COPD subjects referred to the pulmonary clinic of Imam Khomeini Hospital in Sari, Iran.

### Inclusion and Exclusion criteria

The inclusion criteria for the patients were being at the age of over 45 years (owing to higher incidence of COPD in this age group), having COPD, smoking, and being referred by a pulmonologist with airway obstruction diagnosis. In this study, the women did not meet the eligible criteria.

The exclusion criteria were the presence of other systemic medical diseases such as diabetes mellitus, respiratory failure, normal primary spirometry, contraindications for nicotine gum consumption (allergy, recent heart attacks, dangerous arrhythmias, severe angina, hyperthyroidism, insulin-dependent diabetes mellitus, active peptic ulcers, pregnancy, and lactation), and severe psychiatric disorders, such as psychosis, and severe depression and anxiety reported with the patient, and the GSC psychotherapist and psychiatrist diagnosis.

### Randomization, concealment, and blinding

A computerized random number generator was used to produce a randomization schedule employing block randomization (block size of six and twelve) by an independent clinical epidemiologist, who is not involved in the study conducted (recruitment, intervention, and assessment). The randomization sequences were concealed in lightproof, sealed envelopes. Eligible participants were randomly assigned to three parallel groups at a 1:1:1 allocation ratio, according to the randomization list after signing informed consent form. Participants and the therapist were blinded in the initial assessment for inclusion criteria, but, neither participants nor the therapist were blinded during the clinical trial. It was not feasible to mask participants to allocation to GSC or NRT or combined group.

### Interventions

The interventions employed were GSC, NRT and combined of GSC and NRT in three groups.

### NRT Treatment

In this study, NRT was used through transmucosally delivered Nicotine (Nicolife®) blisters (30 labelled gums containing 2 mg nicotine each in an ad lib dose schedule in patients(21). The patients were trained to consume it, and the side effects were explained. NRT was used only in NRT and GSC-NRT groups for 6 weeks, every time the patient had craving.

### GSC Treatment

The patients were guided by the principles of motivation increasing and a 6-page self-change booklet. All the treatments in the three groups were delivered by the same psychotherapist, a trained CBT therapist with more than 15 years of practice. The therapist was trained to give GSC intervention by a psychotherapist and a psychologist at a workshop in 3 days and then treated 5 smokers before the intervention. The treatment sessions in the GSC arm of the intervention were randomly tape-recorded to confirm the process fidelity. In this study, we investigate how to cope with craving in high-risk situations for cigarette smoking.

### GSC intervention protocol

The GSC intervention protocol included 5 sessions, including screening and evaluation, deciding to change, coping with risky situations, identifying different solutions to action, and planning for the future. GSC was applied in 5 one-hour sessions for 5 weeks (22) as follows:

#### Session 1: Screening and evaluation

The questionnaires smoking variables were completed. The motivations to quit smoking were also discussed. The homework involved completing the cast-benefit table for quitting or non-quitting of smoking as well as completing the daily smoking form for different conditions. The patients were asked to contribute to the manual given at the beginning of the sessions.

#### Session 2: Deciding to change

The high-risk situations and smoking stimulants were explained. Withdrawal syndrome and reinforcement of the date for quitting were also expounded. Coping skills and assertiveness were taught. Changing negative thought, body relaxation and respiratory practices were taught.

#### Session 3: Discussing risky situations

The high-risk situations were investigated, and the operational solutions were asked from the patients. The main goal here was to overcome temptation to stay abstinent. The obstacles to overcome were reviewed.

#### Session Four: Identify various solutions to action

The patients retargeted the daily consumption of cigarettes. As the homework, the patients were required to control their consumption status, identify new consumption situations and complete the daily consumption form.

#### Session 5: Steps to the future

The patients were asked to evaluate their commitment and complete the cast-benefit table, again and their improvement was also encouraged. Improvement in life quality and identification of possible difficulties to remain abstinent were also regarded. Finally, a number of phone follow-ups were carried out if necessary (23, 24).

### Outcomes

The primary outcomes included smoking quitting rate and spirometry changes in patients over 6, 12, and 29 weeks after the treatment. The secondary outcomes included (Fagerstrom Test for Nicotine Dependence) FTND and clinical assessment test (CAT) in similar periods in the participants.

### Procedures and measurements

After randomization, additional primary data such as education, smoking quitting history, medical history, and other related data were collected. Cigarette smoking state was measured over 6, 12, and 29 weeks after intervention. Spirometry parameters were analyzed in similar periods. A sample size of 57 participants (19 persons in each group) was considered (conferred 80% power, with two-sided p=0.05) to identify a 10% absolute difference in quitting rates in the three groups.

After explaining the study procedure, the patients completed the demographic information questionnaires and Fagerstrom Test for Nicotine Dependence (FTND) (19, 26–28). The pulmonary function was evaluated based on the results of spirometry (SpiroScout ®, Ganshorn Medizin Electronic, Niederlauer, Germany) (29, 30). FEV1 and FVC were measured by the spirometry device (31, 32) before the intervention and during the treatment to assess further decline/improvement of lung function. The normal spirometry results were defined as FEV1/FVC -70% and FVC -080% (33, 34). In addition to common treatments (e.g., bronchodilator corticosteroid, beta-agonists, and anticholinergic inhalers), NRT and GSC were performed by a trained psychotherapist. NRT was used only in NRT and GSC-NRT groups. After the intervention, the patients were asked to complete the questionnaires again, and the spirometry was repeated over 6, 12, and 29 weeks after the treatment.

### Statistical analysis

To assess the normality of the data distribution, Shapiro-Wilk test was used. The characteristics concerned with the descriptive baseline for the three groups were shown as mean (SD), median (inter-quartile range), or as percentages. Comparison of three groups for categorical data was statistically analyzed through chi-square or Fisher exact test. Additionally, they were statistically analyzed using t-test or Mann-Whitney U test for continuous data. Using intention-to-treat analysis, the primary efficacy data on smoking cessation and pulmonary function were examined. The results in the three groups were analyzed using general linear model (GLM) comparing them in the three groups by repeated measure ANOVA test. The evaluation time was regarded as the within-patient factor. Furthermore, the intervention type (GSC or NRT) was assumed as the between-patient factor. The time groups (interaction term) were regarded as group differences (across the three groups) in their response over time. For compound symmetry assumption, Mauchley’s sphericity test was used. To estimate the differences in values of smoking cessation, pulmonary function at each point in time in the three groups and the time trend after treatment, generalized estimating equation (GEE) was used. A p-value of 0.05 or less was assumed statistically significant. At last, using IBM SPSS statistics version 16 and Stata version 12, the data were analyzed.

## Results

### Participants

In the current study, a total of 900 patients referring to pulmonology clinic were screened. Of the patients, 120 patients declined to participate in the study and 720 patients did not satisfy the eligibility criteria. In this respect, the remaining 60 patients were randomly assigned to three groups of whom three patients were lost to follow-up during the study period. Totally, 57 patients completed the present study and their data were analyzed (Fig. 1).

**Fig 1.**
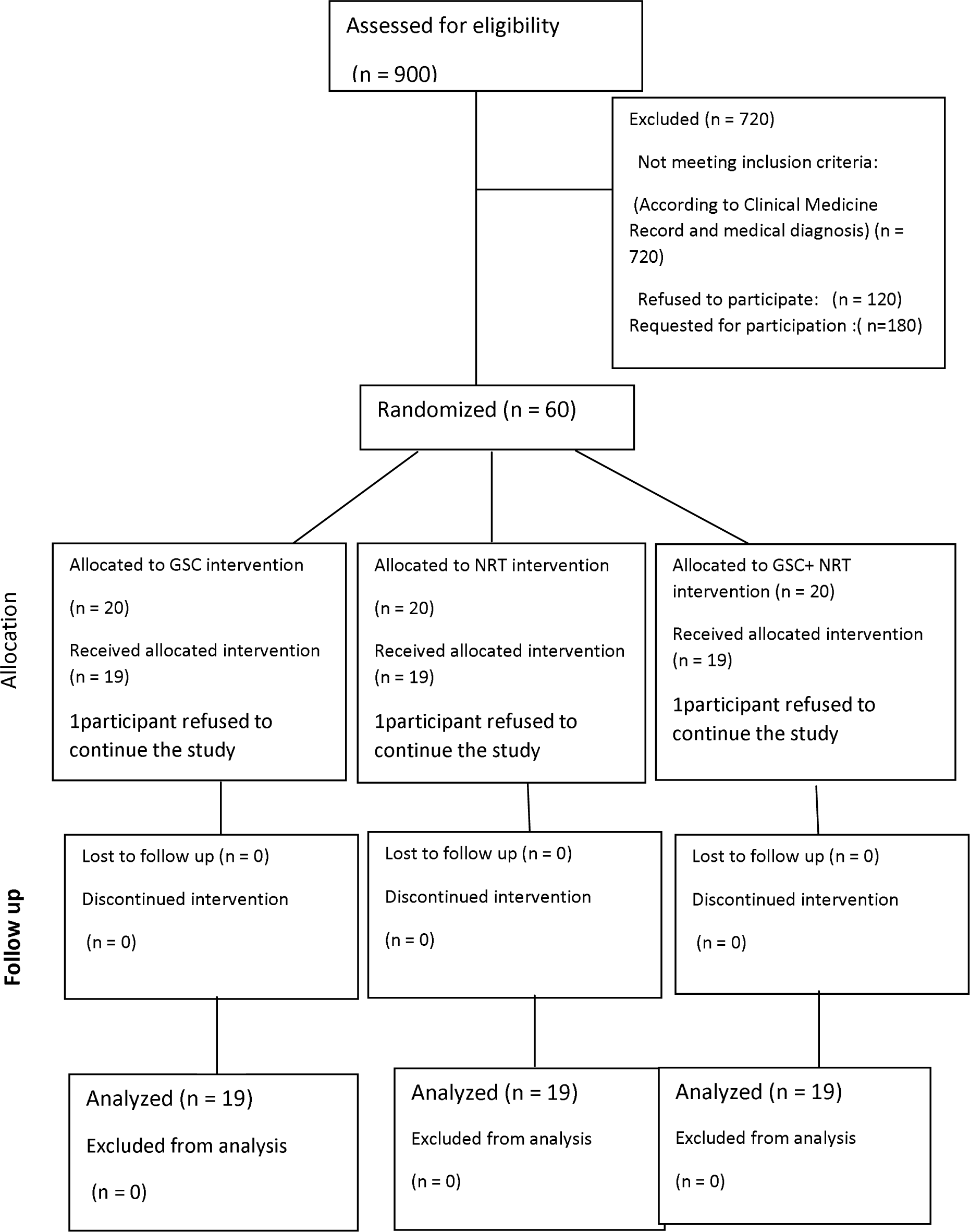
CONSORT diagram of patients’ randomization, intervention, and analysis

Table 1 presents the participants’ clinical characteristics and basic demographic in the three groups. In this study, the women did not meet the eligible criteria, As the table shows, there is no statistically significant difference in average age (GSC vs NRT vs combined group; P=0.08), marital status (GSC vs. NRT vs combined group; P= 0.36), and other characteristics (job, motivation for quitting, importance of smoking cessation, smoker friends, craving, FTND, and daily cigarette smoking). The age of smoking onset was 8-34 (mean=19.6) years and the duration of smoking was 9-59 (mean=32.9) years. A total of 53 patients (93%) smoked after the main meal, 13 (8.22%) regularly smoked after sex; 15 (26.3%) were opium smokers, 4 (7%) took methadone, and 5 (8.8%) also consumed alcohol.

**Table 1:**
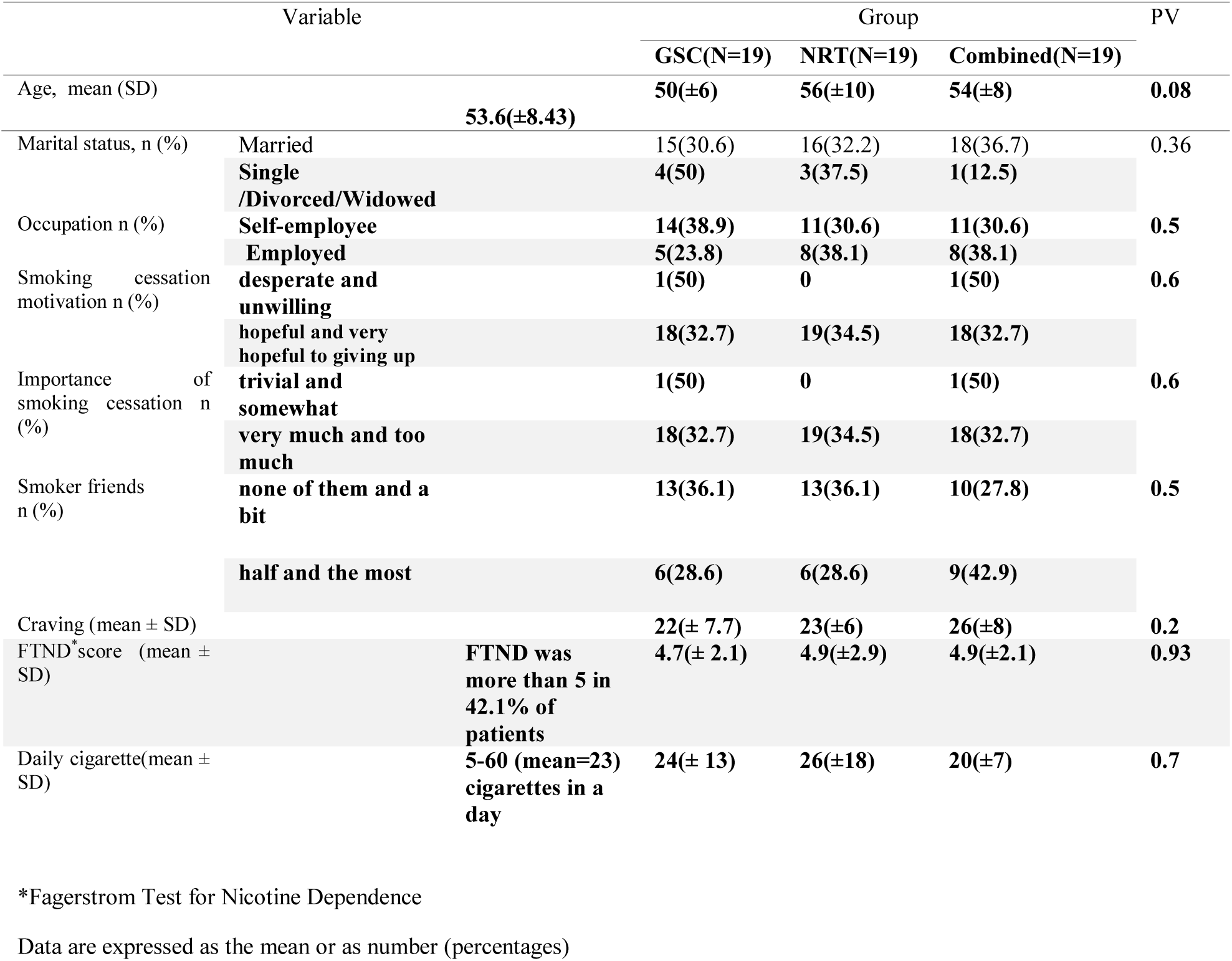
Basic demographic and clinical characteristics of patients in three groups

**Table 2:**
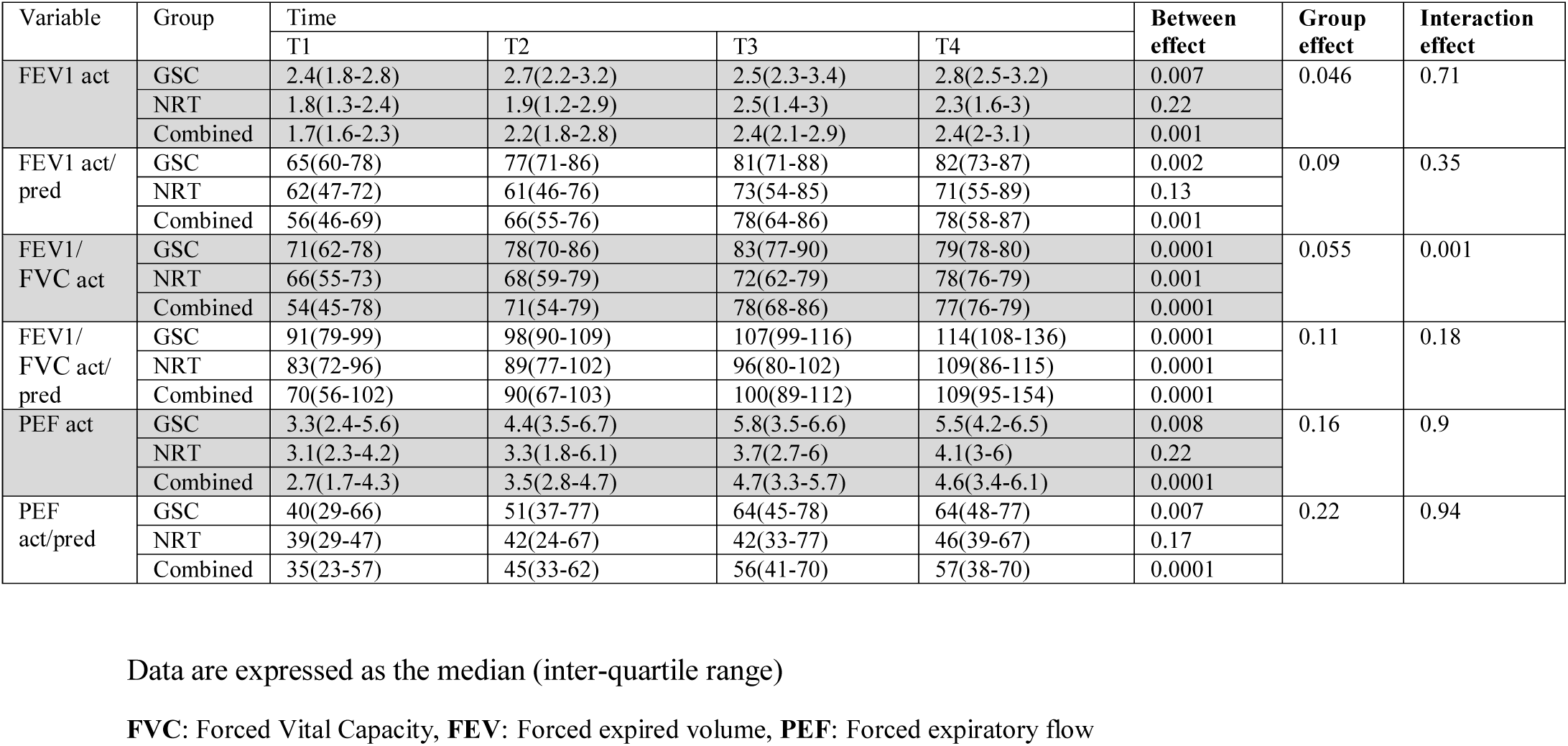
Spirometry characteristics of participants (scores at baseline, 6, 12 and 29 weeks after treatment) in three groups

| Variable | Group | Time |  |  |  | Between effect | Group effect | Interaction effect |
| --- | --- | --- | --- | --- | --- | --- | --- | --- |
|  |  | T1 | T2 | T3 | T4 |  |  |  |
| FEV1 act | GSC | 2.4(1.8-2.8) | 2.7(2.2-3.2) | 2.5(2.3-3.4) | 2.8(2.5-3.2) | 0.007 | 0.046 | 0.71 |
|  | NRT | 1.8(1.3-2.4) | 1.9(1.2-2.9) | 2.5(1.4-3) | 2.3(1.6-3) | 0.22 |  |  |
|  | Combined | 1.7(1.6-2.3) | 2.2(1.8-2.8) | 2.4(2.1-2.9) | 2.4(2-3.1) | 0.001 |  |  |
| FEV1 act/ pred | GSC | 65(60-78) | 77(71-86) | 81(71-88) | 82(73-87) | 0.002 | 0.09 | 0.35 |
|  | NRT | 62(47-72) | 61(46-76) | 73(54-85) | 71(55-89) | 0.13 |  |  |
|  | Combined | 56(46-69) | 66(55-76) | 78(64-86) | 78(58-87) | 0.001 |  |  |
| FEV1/ FVC act | GSC | 71(62-78) | 78(70-86) | 83(77-90) | 79(78-80) | 0.0001 | 0.055 | 0.001 |
|  | NRT | 66(55-73) | 68(59-79) | 72(62-79) | 78(76-79) | 0.001 |  |  |
|  | Combined | 54(45-78) | 71(54-79) | 78(68-86) | 77(76-79) | 0.0001 |  |  |
| FEV1/ FVC act/ pred | GSC | 91(79-99) | 98(90-109) | 107(99-116) | 114(108-136) | 0.0001 | 0.11 | 0.18 |
|  | NRT | 83(72-96) | 89(77-102) | 96(80-102) | 109(86-115) | 0.0001 |  |  |
|  | Combined | 70(56-102) | 90(67-103) | 100(89-112) | 109(95-154) | 0.0001 |  |  |
| PEF act | GSC | 3.3(2.4-5.6) | 4.4(3.5-6.7) | 5.8(3.5-6.6) | 5.5(4.2-6.5) | 0.008 | 0.16 | 0.9 |
|  | NRT | 3.1(2.3-4.2) | 3.3(1.8-6.1) | 3.7(2.7-6) | 4.1(3-6) | 0.22 |  |  |
|  | Combined | 2.7(1.7-4.3) | 3.5(2.8-4.7) | 4.7(3.3-5.7) | 4.6(3.4-6.1) | 0.0001 |  |  |
| PEF act/pred | GSC | 40(29-66) | 51(37-77) | 64(45-78) | 64(48-77) | 0.007 | 0.22 | 0.94 |
|  | NRT | 39(29-47) | 42(24-67) | 42(33-77) | 46(39-67) | 0.17 |  |  |
|  | Combined | 35(23-57) | 45(33-62) | 56(41-70) | 57(38-70) | 0.0001 |  |  |

Data are expressed as the median (inter-quartile range)
**FVC:** Forced Vital Capacity, **FEV:** Forced expired volume, **PEF:** Forced expiratory flow

### Smoking cessation rate

The smoking abstinence rates in NRT, GSC, and combined groups were 21.1% (4 patients), 47.4 %(9 patients), and 47.4 (9 patients), respectively. A statistically difference was significant between the smoking cessation rate in the GSC or combined groups with that in the NRT group (P=0.001). After adjustment of other variables, the GEE model indicated that GSC decreased odds of quitting smoking as compared to those in the NRT group (OR= 0.31, 95% CI: 0.022-0.545).

### Daily cigarette reduction and Nicotine dependence test

The GEE model revealed that daily cigarette reduction in the GSC was higher than that combined group and then in the NRT group, and a statistically significant group effect (P=0.003) was found (table3).

**Table 3:** Daily Cigarette of participants (scores at baseline and time intervals of three weeks in the groups

|  |  | Time |  |  |  |  |  |  |  |  |  | Between effect | Group effect | Interaction effect |
| --- | --- | --- | --- | --- | --- | --- | --- | --- | --- | --- | --- | --- | --- | --- |
|  |  | T1 | T2 | T3 | T4 | T5 | T6 | T7 | T8 | T9 | T10 |  |  |  |
| Daily cigarette | GSC | 20(16-30) | 12(1-18) | 9(1-15) | 7(1-10) | 4(0-10) | 3(0-9) | 3(0-9) | 3(0-9) | 2(0-9) | 2(0-9) | 0.001 | 0.003 | 0.49 |
|  | NRT | 20(12-30) | 13(6-20) | 12(8-20) | 11(7-20) | 10(5-18) | 10(4-17) | 10(4-16) | 10(3-16) | 10(3-16) | 10(3-16) | 0.001 |  |  |
|  | Combined | 20(15-30) | 6(0-14) | 6(0-12) | 5(0-10) | 3(0-8) | 3(0-8) | 2(0-8) | 2(0-7) | 1(0-7) | 1(0-7) | 0.001 |  |  |

The Fagerstrom Test for Nicotine Dependence (FTND) differences between the three groups were not statistically significant (p=0.097). Decreasing FTND score in GSC was more than both the GSC-NRT and NRT groups after 29 weeks.

### Spirometry parameters

The forced vital capacity (FVC-act) and FEV1/FVC act in the three study groups were statistically significant (group effect, P=0.05) (table 2). Contrasts revealed the FVC-act and the FEV1/FVC-act levels in GSC (P=0.03 and P=0.04 respectively) and combined group (P=0.04 and P=0.05 respectively) was higher than NRT group. After adjusting for other variables, GEE model revealed the FVC-act level of the NRT group to be lower than the GSC group (−0.52, 95% CI: -1- -0.02, P=0.04).

The level of FEV_1_-act in the NRT group was lower than in the GSC group (−0.5, 95% CI: -0.9- -0. 12, P=0.009) and it was also lower in the combined group than in the GSC group (−0.38, 95% CI: -0.72- -0.05, P=0.03).The FEV_1_-act/pred level in the NRT group was lower than in the GSC group (−9.7, 95% CI: -17.9- -1.5, P=0.02). The level of FEV1/FVC-act in the NRT group was lower than in the GSC (−5.9, 95% CI: -10.4-(−1.5), P=0.03).Furthermore, this value was also less in the combined group than in the GSC group (−5.9, 95% CI: -10.4-(−1.5), P=0.03). The level of FEV1/FVC act/pred in the NRT group was lower than in the GSC group (−6, 95% CI: -10.4-(- 1.5), P=0.008) and it was also lower in the combined group than in the GSC group (−6, 95% CI: -11.3-(−0.5), PV=0.03) (table 2).

## Discussion

The present study aimed to determine the effectiveness of GSC and NRT on smoking cessation in COPD patients. In this respect, an effective treatment can use pharmacotherapy such as nicotine replacement therapy (NRT) and counseling(35).

A number of studies have been performed into the effect of psychological interventions in smoking cessation. Many pharmaceutical therapies have also been used in the treatment of smoking dependence such as NRT. However, no related study was found on the effects of GSC on the tobacco cessation in COPD patients. To the best of our knowledge, the current study is the first randomized controlled trial demonstrating the efficacy of GSC, NRT, and the combination of them on smoking cessation.

In this study, GSC and combined GSC-NRT were found to be equally effective in tobacco cessation in decreasing smoking. Moreover, serial changes in spirometry results in COPD patients during a smoking cessation trial were examined. Furthermore, the results of our study correlate with previous studies evaluating the effectiveness of counseling, advice, or NRT for smoking cessation in COPD smokers in Iran and other countries. In this regard, cluster randomized controlled trials were carried out with counseling and psychotherapy combined with NRT or bupropion administration, and the usual drug therapy without any psychiatric interventions. The two study groups were treated as one in the analysis since the interventions were found to be equally effective. Additionally, smoking cessation is an effective method to reduce the disease progress by slowing down the decline rate of the annual FEV1 (36). Hilberink SR in a meth analysis evaluated two counseling plans alone or combination with NRT in COPD smokers, the intervention led to a significantly self-reported higher success smoking rate. It also verified quit rates compared to usual care biochemically (37). The interventions resulted in a significantly higher self-reported success rate (14.5%) compared to the usual care (without psychiatric intervention) (7.4%)(37). Also, Lou P et al. obtained 44.3% abstinence rates with behavioral intervention during 48 months (38), Sharifirad GR et al. have shown 46% stable cessation in the group therapy compared to individual counseling(26), Sotoodeh Asl N et al. reported a65.4% reduction of smoking with individual short term CBT (39). In a metha analysis, Lancaster T & Stead LF concluded that, there is a high-quality evidence that individually-delivered smoking cessation counselling can assist smokers to quit (40). Moreover, Sharifi H in an observational study showed that 4.64% of people decreased the number of cigarettes by at least 50%, and 9.12% stopped smoking with behavioral and pharmaceutical therapy (41). Likewise, in our study, GSC and combined GSC-NRT interventions were equally effective in complete abstinence rate. Perhaps, severity of the illness and the the motivation of patients to quit smoking, as well as the therapist’s skills and clinical experience play an important role. Furthermore, the differences between the counseled group and the controls in rates of cessation in 6 months were 33.3% vs 21.4% (42) in a randomized trial of smoking cessation counseling and a self-help manual and three to eight 15-20-min counseling sessions. In a hospital-based randomized smoking cessation trial with counseling and outpatients’ telephone follow-up and minimal counseling and transdermal NRT, 35% of the intensive intervention group reported quitting compared to 21% of the comparison group in 6 months (43). Moreover, evaluation of 21 randomized controlled trials indicated that cognitive therapies combined with medication probably improved smoking abstinence rates, compared to medication (44). One study in Sweden (45) yielded similar results and the smokers received brief smoking cessation advice and annual spirometry, followed by a personal letter from a physician. Three years later, 25% of the smokers with COPD were smoke-free for ≥1 year, compared to 7% of smokers with normal lung function. In addition, a study in Finland indicated that offering individual counseling to all individuals led to a sustained abstinence rate of 5.4% in smokers without COPD, compared to 10.6% in patients with COPD. Nevertheless, success in quitting was not considered to be related to airway obstruction (46). The results of the present study show that GSC is more effective than NRT and NRT-GSC in assisting smoker COPD patients to quit and improve their lung function.

## Conclusion

As a conclusion, GSC smoking cessation counseling can be an effective method to help COPD patients to quit smoking and lead to increased lung health benefits. In this respect, health professionals should offer GSC to smokers in order to help them to quit smoking. Healthy status factors, quitting motivation, and individual counseling are important factors associated with smoking cessation results.

## Data Availability

Data is available when request

## Limitations

A number of limitations existed in the present study. In this study, the most of the patients were from Mazandaran Province in Iran. Therefore, the study population may not correctly represent all smokers in Iran. Moreover, our patients were all men. Since men and women revealed different responses (21), conducting a clinical trial in order to compare the efficacy of GSC in men and women would be a reasonable and interesting study. Furthermore, nicotine dependence and demographic characteristics were not considered to be independent factors for smoking cessation, possibly due to our study small sample size. Moreover, at least, many of our patients consumed opium, alcohol or both of them with their cigarette and interventions was effective in this group, the authors think that these interventions were applied in smokers without these substances.

## Contributors

MZ conceived the original idea for the trial, and sought and obtained the necessary funding. FT, MZ, and ASH designed the study protocol. FT conducted the GSC, managed the day-to-day running of the trial, including all participant follow-up. AA did the statistical analyses. MZ, FT and AA contributed to the clinical interpretation of the results. FT wrote the first draft of the manuscript. MZ, AA, and ASH revised the manuscript critically for important intellectual content. All the authors read and approved the final version.

## Acknowledgements

This paper was originated from the first author’s PhD thesis at Faculty of Medicine, Addiction Research Institute, Mazandaran University of Medical Sciences. The authors thank the participants, research assistants, our colleagues and Mazandaran University of Medical sciences for their in valuable support and care for the study participants.

## References

1. Quist-Paulsen P. Cessation in the use of tobacco - pharmacologic and non-pharmacologic routines in patients. The clinical respiratory journal. 2008;2(1):4–10.

2. Jimenez Ruiz CA, Ramos Pinedo A, Cicero Guerrero A, Mayayo Ulibarri M, Cristobal Fernandez M, Lopez Gonzalez G. Characteristics of COPD smokers and effectiveness and safety of smoking cessation medications. Nicotine & tobacco research: official journal of the Society for Research on Nicotine and Tobacco. 2012;14(9):1035–9.

3. Sundblad BM, Larsson K, Nathell L. High rate of smoking abstinence in COPD patients: Smoking cessation by hospitalization. Nicotine & tobacco research: official journal of the Society for Research on Nicotine and Tobacco. 2008;10(5):883–90.

4. Mulhall P, Criner G. Non-pharmacological treatments for COPD. Respirology (Carlton, Vic). 2016;21 (5):791–809.

5. Adeloye D, Chua S, Lee C, Basquill C, Papana A, Theodoratou E, et al. Global and regional estimates of COPD prevalence: Systematic review and meta-analysis. Journal of global health. 2015;5(2):020415.

6. Lozano R, Naghavi M, Foreman K, Lim S, Shibuya K, Aboyans V, et al. Global and regional mortality from 235 causes of death for 20 age groups in 1990 and 2010: a systematic analysis for the Global Burden of Disease Study 2010. Lancet (London, England). 2012;380(9859):2095–128.

7. Yang IA, Brown JL, George J, Jenkins S, McDonald CF, McDonald VM, et al. COPD-X Australian and New Zealand guidelines for the diagnosis and management of chronic obstructive pulmonary disease: 2017 update. The Medical journal of Australia. 2017;207(10):436–42.

8. Ghobadi H, Ahari SS, Kameli A, Lari SM. The Relationship between COPD Assessment Test (CAT) Scores and Severity of Airflow Obstruction in Stable COPD Patients. Tanaffos. 2012;ll(2):22–6.

9. Kurosawa H. [Clinical examinations for COPD], Rinsho byori The Japanese journal of clinical pathology. 2000;48(12):1118–24.

10. Enright RL, Connett JE, Bailey WC. The FEV1/FEV6 predicts lung function decline in adult smokers. Respiratory medicine. 2002;96(6):444–9.

11. Jimenez-Ruiz CA, Fagerstrom KO. Smoking cessation treatment for COPD smokers: the role of pharmacological interventions. Monaldi archives for chest disease = Archivio Monaldi per le malattie del torace. 2013;79(1):27–32.

12. Tashkin DP. Smoking Cessation in Chronic Obstructive Pulmonary Disease. Seminars in respiratory and critical care medicine. 2015;36(4):491–507.

13. Pezzuto A, Stumbo L, Russano M, Crucitti P, Scarlata S, Caricato M, et al. “Impact of Smoking Cessation Treatment” on Lung Function and Response Rate in EGFR Mutated Patients: A Short-Term Cohort Study. Recent patents on anti-cancer drug discovery. 2015;10(3):342–51.

14. Sobell LC, Sobell MB, Agrawal S. Randomized controlled trial of a cognitive–behavioral motivational intervention in a group versus individual format for substance use disorders. Psychology of Addictive Behaviors. 2009;23(4):672.

15. Sobell MB, Sobell LC. Guided self-change model of treatment for substance use disorders. Journal of Cognitive Psychotherapy. 2005;19(3):199–210.

16. Saladin ME, Santa Ana EJ. Controlled drinking: More than just a controversy. Current Opinion in Psychiatry. 2004;17(3):175–87.

17. Wilson GT, Zandberg LI. Cognitive-behavioral guided self-help for eating disorders: effectiveness and scalability. Clinical psychology review. 2012;32(4):343–57.

18. Riaz M, Lewis S, Coleman T, Aveyard P. Which measures of cigarette dependence are predictors of smoking cessation during pregnancy? Analysis of data from a randomized controlled trial. 2016;111(9):1656–65.

19. Sarbandi F, Niknami S, Hidarnia A, Hajizadeh E, Masooleh HA, Nobari SE. Psychometric properties of the Iranian version of the fagerstrom test for nicotine dependence and of heaviness of smoking index. Journal of Research & Health. 2015;5(1):96–103.

20. Velicer WF, DiClemente CC, Prochaska JO, Brandenburg N. Decisional balance measure for assessing and predicting smoking status. Journal of personality and social psychology. 1985;48(5):1279.

21. Killen. Evaluation of a treatment approach combining nicotine gum with self-guided behavioral treatments for smoking relapse prevention. J Consult Clin Psychol. 1990;58(1):85–92.

22. Camarelles F, Asensio A, Jiménez-Ruiz C, Becerril B, Rodero D, Vidaller O. [Effectiveness of a group therapy intervention to quit smoking. Randomized clinical trial]. Medicina clínica. 2002;119(2):53–7.

23. Sobell LC, Sobell MB, Agrawal S. Randomized controlled trial of a cognitive-behavioral motivational intervention in a group versus individual format for substance use disorders. Psychology of addictive behaviors: journal of the Society of Psychologists in Addictive Behaviors. 2009;23(4):672–83.

24. Guimarães FMCL, Nardi AE, Cardoso A, Valença AM, Conceição EGd, King ALSJM. Cognitive behavioral therapy treatment for smoking alcoholics in outpatients. 2014;1(6):336–40.

25. Chuang ML, Lin IF, Lee CY. Clinical assessment tests in evaluating patients with chronic obstructive pulmonary disease: A cross-sectional study. Medicine. 2016;95(47):e5471.

26. Sharifirad GR, Eslami AA, Charkazi A, Mostafavi F, Shahnazi H. The effect of individual counseling, line follow-up, and free nicotine replacement therapy on smoking cessation in the samples of Iranian smokers: Examination of transtheoretical model. Journal of research in medical sciences: the official journal of Isfahan University of Medical Sciences. 2012;17(12):1128.

27. Masjedi MR, Azaripour Masooleh H, Hosseini M, Heydari GR. Effective factors on smoking cessation among the smokers in the first “smoking cessation clinic” in Iran. Tanaffos. 2002;1(4):61–7.

28. Heydari GR, Ariyanpour M, Kashani BS, Ramezankhani A, Tafti SF, Hosseini M, et al. Tobacco dependency evaluation with fagerstrom test among the entrants of smoking cessation clinic. Tanaffos. 2007;6(4):47–52.

29. Rigotti NA. Smoking cessation in patients with respiratory disease: existing treatments and future directions. The Lancet Respiratory Medicine. 1(3):241–50.

30. Vestbo J, Hurd SS, Agusti AG, Jones PW, Vogelmeier C, Anzueto A, et al. Global strategy for the diagnosis, management, and prevention of chronic obstructive pulmonary disease: GOLD executive summary. American journal of respiratory and critical care medicine. 2013;187(4):347–65.

31. BTS guidelines for the management of chronic obstructive pulmonary disease. The COPD Guidelines Group of the Standards of Care Committee of the BTS. Thorax. 1997;52 Suppl 5:S1-28.

32. Miravitlles M S-CJ, Calle M, Molina J, Almagro P, et al. (2012) Pharmacological treatment of stable COPD. Arch Bronconeumol 48: 247–257.

33. Simmons MS, Connett JE, Nides MA, Lindgren PG, Kleerup EC, Murray RP, et al. Smoking reduction and the rate of decline in FEV(1): results from the Lung Health Study. The European respiratory journal. 2005;25(6):1011–7.

34. Guerra S, Carsin AE, Keidel D. Health-related quality of life and risk factors associated with spirometric restriction. 2017;49(5).

35. Heydari G, Masjedi M, Ahmady AE, Leischow SJ, Lando HA, Shadmehr MB, et al. A comparative study on tobacco cessation methods: a quantitative systematic review. International journal of preventive medicine. 2014;5(6):673–8.

36. Drake P, Driscoll AK, Mathews TJ. Cigarette Smoking During Pregnancy: United States, 2016. NCHS data brief. 2018(305):1-8.

37. Hilberink SR, Jacobs JE, Breteler MH, de Vries H, Grol RP. General practice counseling for patients with chronic obstructive pulmonary disease to quit smoking: impact after 1 year of two complex interventions. Patient education and counseling. 2011;83(1):120–4.

38. Lou P, Zhu Y, Chen P, Zhang P, Yu J, Zhang N, et al. Supporting smoking cessation in chronic obstructive pulmonary disease with behavioral intervention: a randomized controlled trial. BMC family practice. 2013;14:91.

39. Sotoodeh Asl N, Taher Neshatdost H, Kalantari M, Talebi H, Mehrabi HA, Khosravi AR. The effectiveness of cognitive behavioral therapy on the reduction of Tobacco Dependency in patients with essential hypertension. Journal of Research in Behavioural Sciences. 2011;9(2):94–103.

40. Lancaster T, Stead LF. Individual behavioural counselling for smoking cessation. Cochrane Database of Systematic Reviews. 2017(3).

41. Sharifi H, Kharaghani R, Emami H, Hessami Z, Masjedi MR. Efficacy of harm reduction programs among patients of a smoking cessation clinic in Tehran, Iran. Arch Iran Med. 2012;15(5):283–9.

42. Pederson LL, Wanklin JM, Lefcoe NM. The effects of counseling on smoking cessation among patients hospitalized with chronic obstructive pulmonary disease: a randomized clinical trial. The International journal of the addictions. 1991;26(1):107–19.

43. Simon JA, Carmody TP, Hudes ES, Snyder E, Murray J. Intensive smoking cessation counseling versus minimal counseling among hospitalized smokers treated with transdermal nicotine replacement: a randomized trial. The American journal of medicine. 2003;114(7):555–62.

44. Denison E, Underland V, Mosdol A, Vist G. NIPH Systematic Reviews. Cognitive Therapies for Smoking Cessation: A Systematic Review. Oslo, Norway: Knowledge Centre for the Health Services at The Norwegian Institute of Public Health (NIPH) Copyright (c) 2017 by The Norwegian Institute of Public Health (NIPH). 2017.

45. Stratelis G, Molstad S, Jakobsson P, Zetterstrom O. The impact of repeated spirometry and smoking cessation advice on smokers with mild COPD. Scand J Prim Health Care. 2006;24(3):133–9.

46. Toljamo T, Kaukonen M, Nieminen P, Kinnula VL. Early detection of COPD combined with individualized counselling for smoking cessation: a two-year prospective study. Scand J Prim Health Care. 2010;28(1):41–6.

